# Analytical and clinical performances of a SARS-CoV-2 S-RBD IgG assay: comparison with neutralization titers

**DOI:** 10.1101/2021.03.10.21253260

**Authors:** Andrea Padoan, Francesco Bonfante, Chiara Cosma, Costanza Di Chiara, Laura Sciacovelli, Matteo Pagliari, Alessio Bortolami, Paola Costenaro, Giulia Musso, Daniela Basso, Carlo Giaquinto, Mario Plebani

**Author notes:** **Corresponding Author**: Prof. Mario Plebani, MD, Department of Medicine-DIMED, University of Padova and Department of Laboratory Medicine, University-Hospital of Padova (Italy), Via Giustiniani 2, 35128, Padova, Italy.

## Abstract

**Background:** SARS-CoV-2 serology presents an important role in understanding the virus epidemiology, in vaccine prioritization strategies and in convalescent plasma therapy. Immunoassays performances have to be accurately evaluated and correlated with neutralizing antibodies to be used as a surrogate measure of neutralizing activity. We investigate the analytical and clinical performance of a SARS-CoV-2 RBD IgG assay, automated on a high throughput platform, and the correlation of the antibodies (Ab) levels with the plaque reduction neutralization (PRNT_50_) Ab titers.

**Methods:** A series of 546 samples were evaluated by SARS-CoV-2 RBD IgG assay (Snibe diagnostics), including 171 negative and 168 positive SARS-CoV-2 subjects and a further group of 207 subjects of the COVID-19 family clusters follow-up cohort.

**Results:** Assay precision was acceptable at low and medium levels; linearity was excellent in all the measurement range. Considering specimens collected after 14 days post symptoms onset, overall sensitivity and specificity were 99.0% and 92.5%, respectively. A total of 281 leftover samples results of the PRNT_50_ test were available. An elevated correlation was obtained between the SARS-CoV-2 RBD IgG assay and the PRNT_50_ titer at univariate (rho = 0.689) and multivariate (rho = 0.712) analyses.

**Conclusions:** SARS-CoV-2 S-RBD IgG assay achieves elevated analytical and clinical performances, and a strong correlation with sera neutralization activity.

## 1. Introduction

Current testing for SARS-CoV-2 largely depends on labor-intensive molecular techniques, particularly reverse transcription real-time polymerase chain reaction (rRT-PCR), but a body of evidence highlights that individuals with positive molecular tests represent only a small fraction of all infections, given limited availability and the brief time window when rRT-PCR testing presents the highest sensitivity (1,2).

Serological assays for the accurate measurement of SARS-CoV-2 antibodies are suboptimal tools for the early diagnosis of infection but provide important population-based data on pathogen exposure, on the prevalence of infection, also in asymptomatic subjects, and on the selection of convalescent plasma donors. Furthermore, SARS-CoV-2 serology represents a complementary tool of molecular virological assays to achieve a more accurate diagnosis in some “difficult” patients, for tracking transmission dynamics, gaining knowledge on population immunity levels and informing disease control policies (3). In addition, serology plays a central role in clinical trials on vaccine development to provide evidence of potency and efficacy (4,5) and in supporting decisions on population groups who should be prioritized in vaccine administration (6). Many assays have been developed for SARS-CoV-2 antibody detection, including lateral flow tests, enzyme-linked immunosorbent assays (ELISA), chemiluminescent (CLIA) assays and other platforms (https://www.fda.gov/medical-devices/emergency-use-authorizations-medical-devices/coronavirus-disease-2019-covid-19-emergency-use-authorizations-medical-devices); some rely on whole inactivated virions, while others adopted viral subunits such as the nucleocapsid protein, or viral spike protein. Irrespective of the numbers of papers dealing with specificity and sensitivity of these assays, key issues such as the correlation between circulating antibodies and their neutralizing ability and persistence over time have not been adequately addressed. More recently, a body of evidence has been collected to demonstrate that the recombinant SARS-CoV-2 receptor binding domain (RBD) is a highly sensitive and specific antigen for the detection of antibodies induced by SARS-CoV-2 and that the levels of RBD-binding antibodies present a strong correlation with neutralizing antibodies in COVID-19 patients (7,8).

Aim of this paper is the analytical and clinical evaluation of a SARS-CoV-2 RBD IgG assay, automated on a high throughput platform and the correlation of IgG levels with neutralizing antibodies.

## 2. Material and Methods

### 2.1 Patients

A total of 546 leftover serum samples from 168 COVID-19 patients [24 asymptomatic or with /Mild disease (Asympt/Mild), who recovered at home with supportive care and isolation, and 144 hospitalized classified with mild or moderate/severe disease, following WHO interim guidance (9)] and 171 SARS-CoV-2 negative subjects [97 pre-pandemic samples, 31 healthcare workers, 11 and 32 patients with rheumatic disease or with human immunodeficiency virus (HIV)] were included in the study. A further group of 207 subjects of the COVID-19 Family Cluster Follow-up Clinic (CovFC), set up at the Department of Women’s and Children’s Health of the University Hospital of Padua, in the Veneto Region, Italy were studied. Families were enrolled when complied with the following inclusion criteria: a) having children of pediatric age; b) having a history of medically confirmed COVID-19 or being a household member of a COVID-19 confirmed case. All subjects underwent at least one nasopharyngeal swab test analyzed by rRT-PCR. Healthcare workers were considered negative (HCW) on the basis of at least three negative sequential molecular test results obtained between February 26th and May 29th, 2020. Considering family clusters, information concerning their past and recent history were collected retrospectively through both patients interviews and the revision of clinical files. Family subjects who had tested positive for SARS-CoV-2 by RRT-PCR and/or by either of the two serological tests adopted in this study were considered confirmed COVID-19 cases. For each confirmed COVID-19 case, a baseline was defined as the most likely onset of infection, based on different criteria. In detail, for patients reporting COVID-19 related symptoms, the baseline coincided with the onset of symptoms; in case of asymptomatic patients the baseline referred to the date the first positive NP swab was recorded. Among SARS-SARS-CoV-2 patients in family clusters, 5 were hospitalized for moderate disease, whereas the others were recovered at home.

Only authorized staff involved in data entry were provided with passwords for protected access to collected data. All data were collected maintaining confidentiality and were anonymized for statistical analysis. The study protocol (number 23307) was approved by the Ethics Committee of the University-Hospital, Padova. All the patients were informed of the study and voluntarily agreed to participate, providing a written consent.

### 2.2 Analytical system under evaluation

In this study, a commercially available immunoassay was evaluated, the anti-SARS-CoV-2 S-RBD IgG (Snibe Diagnostics, New Industries Biomedical Engineering Co., Ltd [Snibe], Shenzhen, China). SARS-CoV-2 S-RBD IgG is a chemiluminescent immunoassay (CLIA) that determines IgG Ab against the RBD of the Spike (S) protein of the virus, in human serum or plasma. All analyses were performed on MAGLUMI™ 2000 Plus (Snibe Diagnostics), with results expressed in kAU/L.

### 2.3 Repeatability and intermediate precision evaluation

Precision estimation was performed using three human serum sample pools with different values, by means of triplicate measurements of same pool aliquots, performed for a total of five consecutive days. Nested analysis of variance (ANOVA) was used to estimate precision, following the CLSI EP15-A3 protocol (10). The results for precision were compared to those claimed by the manufacturer, using the procedure recommended by EP15-A3.

### 2.4 Linearity assessment of Maglumi anti-SARS-CoV-2 S-RBD

Linearity was assessed using two samples pools (high level pools), prepared with different levels of SARS-CoV-2 antibodies, serially diluted in low level pools, as specified in the CLSI EP06-A guideline (paragraph 4.3.1) (11). The following high-level serum pools were prepared: 3.7 kAU/L and 71 kAU/L. The pools were serially diluted with the corresponding low-level serum pools (0.181 kAU/L and 0.59 kAU/L). All measurements were performed in triplicate. Polynomial regression was used to test deviation from linearity.

### 2.5 Plaque reduction neutralization test (PRNT)

A high-throughput PRNT method was used for the fast and accurate quantification of neutralizing antibodies in plasma samples collected from patients exposed to SARS-CoV-2, as described elsewhere (12). Briefly, after heat-inactivation, samples were diluted in Dulbecco modified Eagle medium (DMEM) and then mixed with a virus solution containing 20-25 focus-forming units (FFUs) of SARS-CoV-2. After 1 hour at 37 °C, fifty microliters of the virus–serum mixtures were added to confluent monolayers of Vero E6 cells, in 96-wells plates and incubated for 1 hour at 37 °C, in a 5% CO_2_ incubator. After 26 h of incubation and cells fixing, visualization of plaques was obtained with an immunocytochemical staining method using an anti-dsRNA monoclonal antibody (J2, 1:10,000; Scicons) for 1 hour, followed by 1-hour incubation with peroxidase-labeled goat anti-mouse antibodies (1:1000; DAKO) and a 7-minute incubation with the True Blue™ (KPL) peroxidase substrate. FFUs were counted after acquisition of pictures at a high resolution of 4800 x 9400 dpi, on a flatbed scanner. The serum neutralization titer was defined as the reciprocal of the highest dilution resulting in a reduction of the control plaque count >50% (PRNT_50_). From previous experiments, we defined a titer of 1:10 as the seropositive threshold (12).

### 2.6 Statistical analyses

For evaluation of precision, an in-house developed R (R Foundation for Statistical Computing, Vienna, Austria) script for implementing the CLSI EP15-A3 protocol was used for ANOVA and for calculating the upper verification limit (10). The GraphPad Prism version 9.1 for Windows (GraphPad Software, LLC) was employed to evaluate plaque reduction neutralization test results. Stata v16.1 (Statacorp, Lakeway Drive, TX, USA) was used to evaluate the assays’ clinical performances. Bonferroni’s adjusted p-value (B-adj) was calculated for multiple comparisons. For ROC analyses, the non-parametric empirical method was used to estimate the area under the ROC curve (AUC), while the ‘diagt’ module was used to estimate sensitivity, specificity, and positive and negative predictive values. Youden index (calculated as *sensitivity+specificity-1*) was used to estimate the best performances of the assay. Considering a type I error α = 0.05, a power of 0.8 and with 249 positive and 249 negative subjects, a sensitivity (or specificity) of 0.95 can be considered significant with respect to values above or equal to 0.99 (null hypothesis). PASS 2020 Power Analysis and Sample Size Software (2020), NCSS, Kaysville, Utah, USA, was used for sample size and power analyses.

## 3. Results

### 3.1 Patients’ characteristics

Demographic characteristics of the subjects included in the study are reported in Table 1. The overall mean age was 42.5 years, with a standard deviation (±SD) of 22.6 (range 0.7 - 92.2 years). Excluding family clusters, the remaining subjects (n=337) presented a mean age (±SD) of 53.7±16.9 years. A multivariate ANOVA analysis was performed considering Age as dependent variable and Gender and studied groups (F=56.55, p < 0.001) as independent variables. The ages of family clusters differed from other groups [Bonferroni’s adjusted (B-adj) p < 0.001 for all], except for Asympt/Mild positive patients [B-adj p = 0.051]. Age of negative healthcare workers (HCW), pre-pandemic subjects and Asympt/Mild patients were not statistically significant different (p = 0.999), while these groups’ age differ with respect to hospitalized COVID patients [B-adj p < 0.0001]. Age of Rheumatic disease/HIV patients differs from other groups (B-adj p < 0.001 for all), with the exception of Asympt/Mild disease group (B-adj p =0.493). The percentage of females differed significantly from that of males (p< 0.001), particularly in the Asympt/Mild disease group. For SARS-CoV-2 patients, the mean time interval from the onset of symptoms and serological determinations was 17.7 days (SD ±16.3; range 1 - 103 days). In the family clusters, the mean time interval from the onset of symptoms and serological determinations was 148.2 days (SD ±71.2; range 41-492 days). The differences in time from symptoms onset with respect to the studied groups of individuals were reported in Table 2.

**Table 1:** Demographic characteristics of the two Cohorts of subjects included in the study.

| Cohorts | Types of individuals | N (%) | Gender |  | Age (mean±SD) |
| --- | --- | --- | --- | --- | --- |
|  |  |  | Females N (%) | Males N (%) |  |
| Group 1 | Pre-Pandemic | 97 (28.6%) | 62 (63.9) | 35 (36.1%) | 40.8±11.9 |
|  | Negative Healthcare Workers (HCW) | 31 (9.1%) | 10 (32.3%) | 21 (67.7%) | 43.6±12.0 |
|  | Patients with rheumatic diseases and with human immunodeficiency virus (AI/HIV) | 43 (12.7%) | 12 (27.9%) | 31 (72.1%) | 52.2±12.7 |
|  | Asymptomatic/Mild SARS-CoV-2 positive Patients (Asympt/Mild) | 24 (7.1%) | 21 (87.5%) | 3 (12.5%) | 43.0±13.9 |
|  | Severe SARS-CoV-2 positive Hospitalized patients (Sev) | 70 (20.6%) | 24 (34.3%) | 46 (65.7%) | 61.3±16.1 |
|  | Critical SARS-CoV-2 positive Hospitalized patients (Critical) | 74 (21.9%) | 14 (18.9%) | 60 (81.1%) | 68.1±13.8 |
|  | All samples of Cohort 1 | 339 (100%) | 143 (42.2%) | 196 (73.8%) | 53.7±16.9 |
| Group 2 | Families with COVID-19 pediatric patients | 207 (37.9%) | 95 (45.9%) | 112 (54.1%) | 24.2±18.4 |
| Overall | All samples | 546 (100%) | 238 (43.6%) | 308 (56.4%) | 42.5±22.6 |

**Table 2:** Disease severity, time from symptoms onset, percentage of positive samples to serological determination of anti-SARS-CoV-2 RBD IgG antibodies and PRNT_50_ titers, subdivided by the studied groups.

| Types of individuals | Samples evaluated for SARS-CoV-2 antibodies (n) | Days from symptoms onset and serology (mean±SD) | Percentage of samples with positive assays results (n, %) | Samples tested for neutralization activity (n) | Percentage of samples with neutralizing antibodies (PRNT<sub>50</sub> ≥ 1:10) |
| --- | --- | --- | --- | --- | --- |
| **Pre-Pandemic** | 97 (28.6%) | - | 4 | 25 | 0 |
| **Negative Healthcare Workers (HCW)** | 31 (9.1%) | - | 8 | - | - |
| **Patients with rheumatic diseases and with human immunodeficiency virus (AI/HIV)** | 43 (12.7%) | - | 2 | - | - |
| **Asymptomatic/Mild SARS-CoV-2 positive Patients (Asympt/Mild)** | 24 (7.1%) | 73±28.3* | 22 (91.6%) | 5 | 5 (100%) |
| **Severe SARS-CoV-2 positive Hospitalized patients (Sev)** | 70 (20.6%) | 16.1±17.5 | 60 (85.7%) | 23 | 23 (100%) |
| **Critical SARS-CoV-2 positive Hospitalized patients (Critical)** | 74 (21.9%) | 19.3±15.0 | 70 (94.6%) | 26 | 26 (100%) |
| **Families with COVID-19 pediatric patients** | 207 (37.9%) | 148.2±71.2§ | 191(92.2%) | 207 | 189 (91.3%) |
| **Overall** | 546 | 89.5±84.2 | 194 | 281 | 238 |

\* data available for only 2 patients; § statistically significant with respect to the time from symptom onset of Severe and Critical hospitalized patients (p < 0.001) (One-way ANOVA, F = 227.7, p < 0.0001)

### 3.2 Repeatability and intermediate precision

Results for precision of CLIA assay is reported in Table 3. Repeatability and within-laboratory precision were in accordance with the repeatability and intermediate precision conditions specified in the international vocabulary of metrology (VIM, JCGM 100:2012) for precision estimation within a five-day period. Obtained data show acceptable imprecision at low and medium levels, but significantly deviated from the values claimed by the manufacturer for the high-level control material.

**Table 3:**
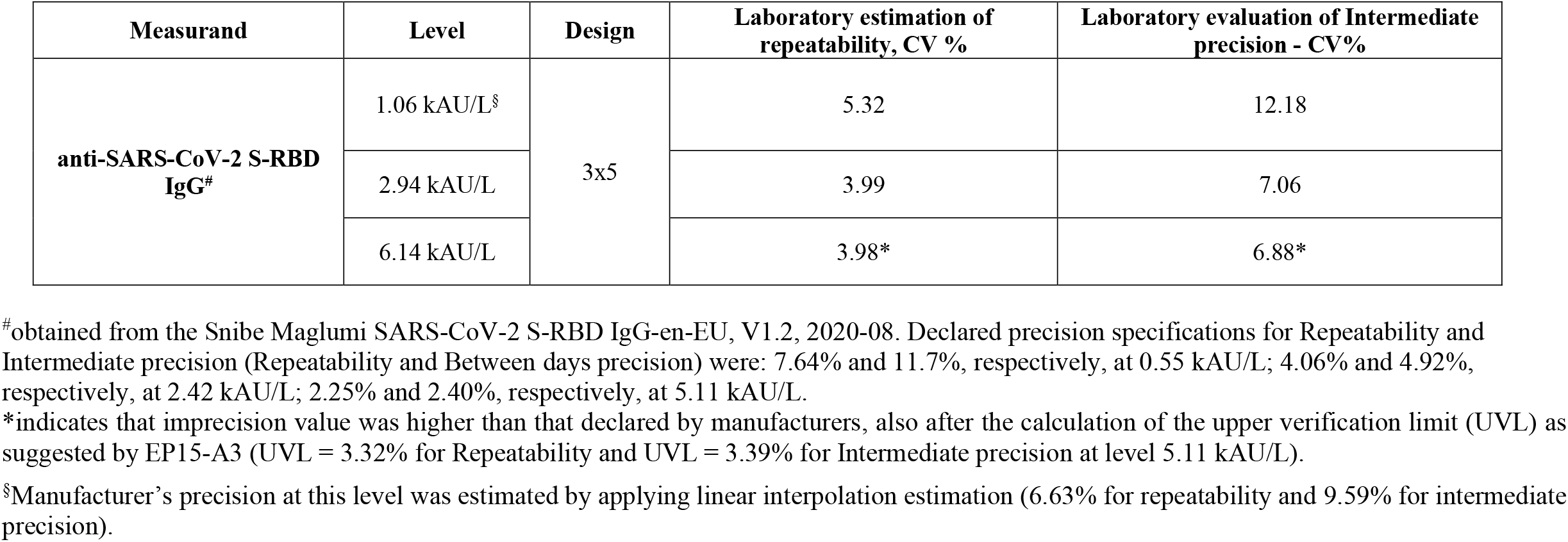
Precision results of Maglumi SARS-CoV-2 S-RBD IgG obtained using a 3×5 design (triplicate measurement for 5 consecutive days). Coefficient of variation (CV) are expressed in percentage (%) and were obtained by using pools of samples.

| Measurand | Level | Design | Laboratory estimation of repeatability, CV % | Laboratory evaluation of Intermediate precision - CV% |
| --- | --- | --- | --- | --- |
| **anti-SARS-CoV-2 S-RBD IgG<sup>#</sup>** | 1.06 kAU/L<sup>§</sup> | 3x5 | 5.32 | 12.18 |
|  | 2.94 kAU/L |  | 3.99 | 7.06 |
|  | 6.14 kAU/L |  | 3.98* | 6.88* |

<sup>#</sup>obtained from the Snibe Maglumi SARS-CoV-2 S-RBD IgG-en-EU, V1.2, 2020-08. Declared precision specifications for Repeatability and Intermediate precision (Repeatability and Between days precision) were: 7.64% and 11.7%, respectively, at 0.55 kAU/L; 4.06% and 4.92%, respectively, at 2.42 kAU/L; 2.25% and 2.40%, respectively, at 5.11 kAU/L.
\*indicates that imprecision value was higher than that declared by manufacturers, also after the calculation of the upper verification limit (UVL) as suggested by EP15-A3 (UVL = 3.32% for Repeatability and UVL = 3.39% for Intermediate precision at level 5.11 kAU/L).
<sup>§</sup>Manufacturer's precision at this level was estimated by applying linear interpolation estimation (6.63% for repeatability and 9.59% for intermediate precision).

### 3.3 Linearity assessment

Linearity results for Maglumi anti-SARS-CoV-2 S-RBD are summarized in Figure 1. Since the method is claimed to be quantitative, tested mixes were prepared for covering a wide range of values (the upper limit of the method without sample dilution is 100 kAU/L), including the manufacturers’ cut-offs. Maglumi anti-SARS-CoV-2 S-RBD IgG does not deviate from linearity in the entire range of tested values, being the coefficients of the second-order polynomial non-statistically significant.

**Figure 1:**
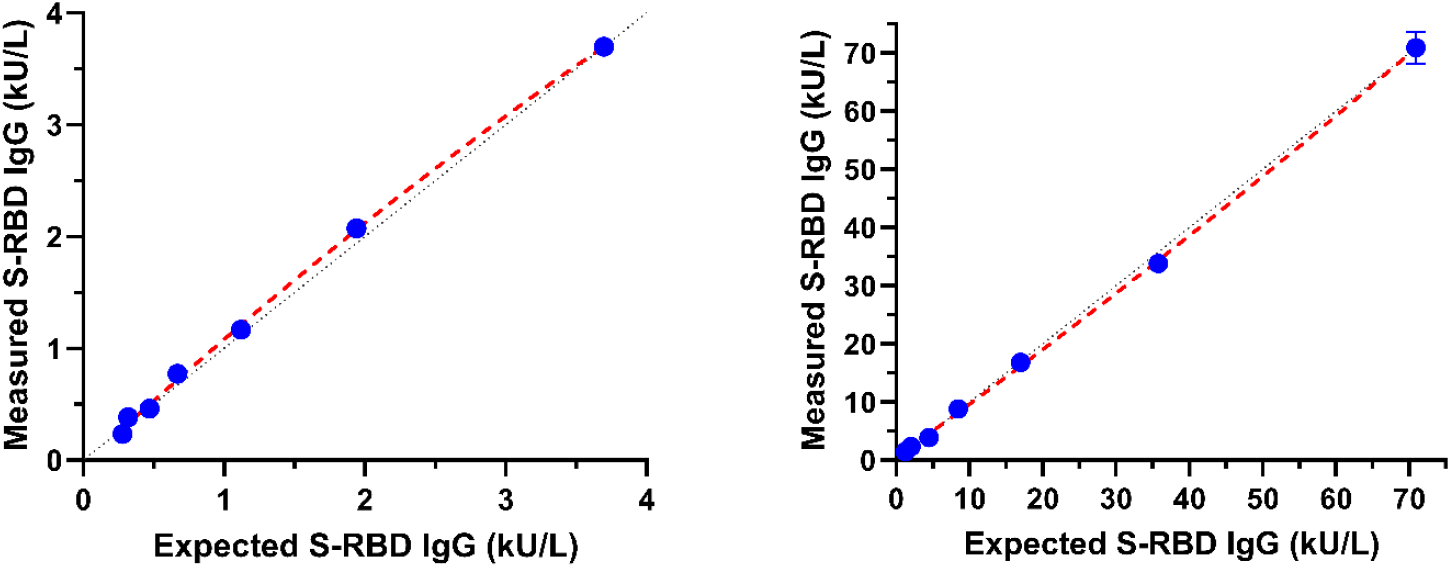
Linearity assessment of anti-SARS-CoV-2 S-RBD IgG assays, performed at two concentration levels.

### 3.4 Evaluation of clinical performances

For a total of 339 samples, including pre-pandemic (collected in 2015), negative HCW and AI/HIV subjects (collected between March 2020 and May 2020) and samples from patients hospitalized for COVID-19 (collected between April 2020 and November 2020), a total of 178 and 161 resulted negative and positive to SARS-CoV-2, respectively. In family clusters of COVID-19, out of 207 samples, 191 had a laboratory-confirmed past SARS-CoV-2 infection, and positivity were correctly identified by the assay in all cases under evaluation.

The distribution of log_10_ anti-SARS-CoV-2 S-RBD transformed results is reported in Figure 2, considering both overall individuals and only samples collected after 14 days post-symptoms onset. Considering only samples collected after 14 days post-symptoms onset, median and interquartile range (IQR) of anti-SARS-CoV-2 S-RBD Ab in SARS-CoV-2 patients were: 18.5 kAU/L (12.13 - 30.48 kAU/L) for Asympt/Mild, 52.1 kAU/L (34.1-78.0 kAU/L) for Severe and 79.1 (36.3 - 100 kAU/L) for Critical individuals; for family Clusters the median and IQR Ab level was 27.3 kAU/L (10.9 - 51.6 kAU/L). By using the Kruskal-Wallis test, significant differences were obtained comparing Asympt/Mild with Severe or Critical patients [Bonferroni’s adjusted (Badj) p-value < 0.001 for both], and between Severe or Critical patients with family clusters [Bonferroni’s adjusted (Badj) p-value < 0.001 for both]; no statistical significance difference was observable between Severe and Critical SARS-CoV-2 patients (Badj p-value = 0.117).

**Figure 2:**
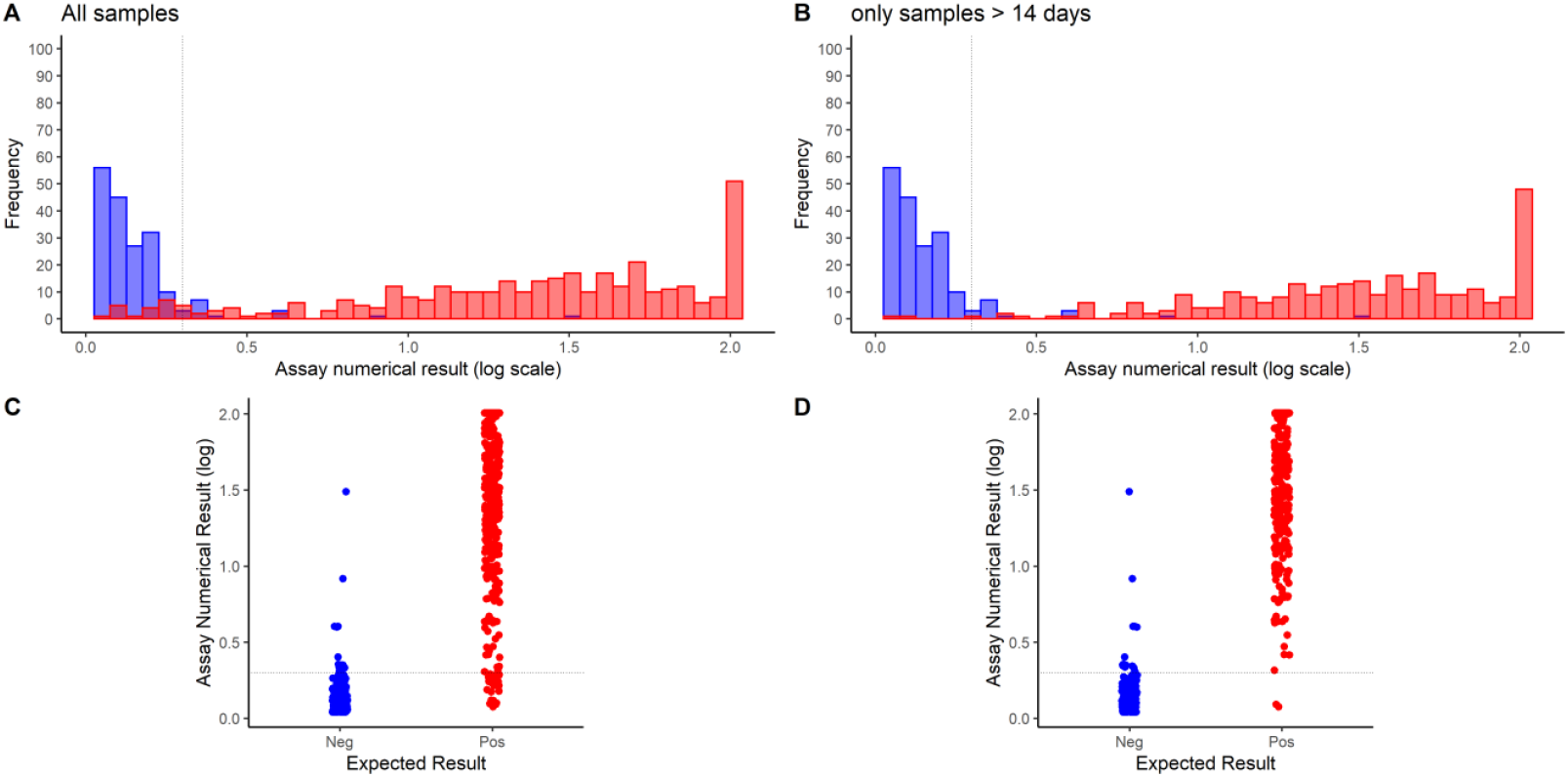
Frequency histograms and dot plots of log_10_ transformed anti-SARS-CoV-2 S-RBD IgG CLIA results (in kAU/L), considering all the studied individuals (A and C) and only samples collected after 14 days from symptom onset (B and D).

Sensitivities, specificities, and positive/negative likelihood ratios, estimated using the manufacturers’ cut-offs and considering samples collected from 14 days post-symptoms onset, were reported in Figure 3 and Supplementary Table 1. Receiver operating characteristic (ROC) curves were further reported in the same table. A further analysis was performed, using the Youden index strategy, for identifying the most accurate cut-off; However, the cut-off calculated with Youden’s index (0.96 kAU/L) does not significantly improve the clinical performances when compared to that recommended by the manufacturer (1.0).

**Figure 3:**
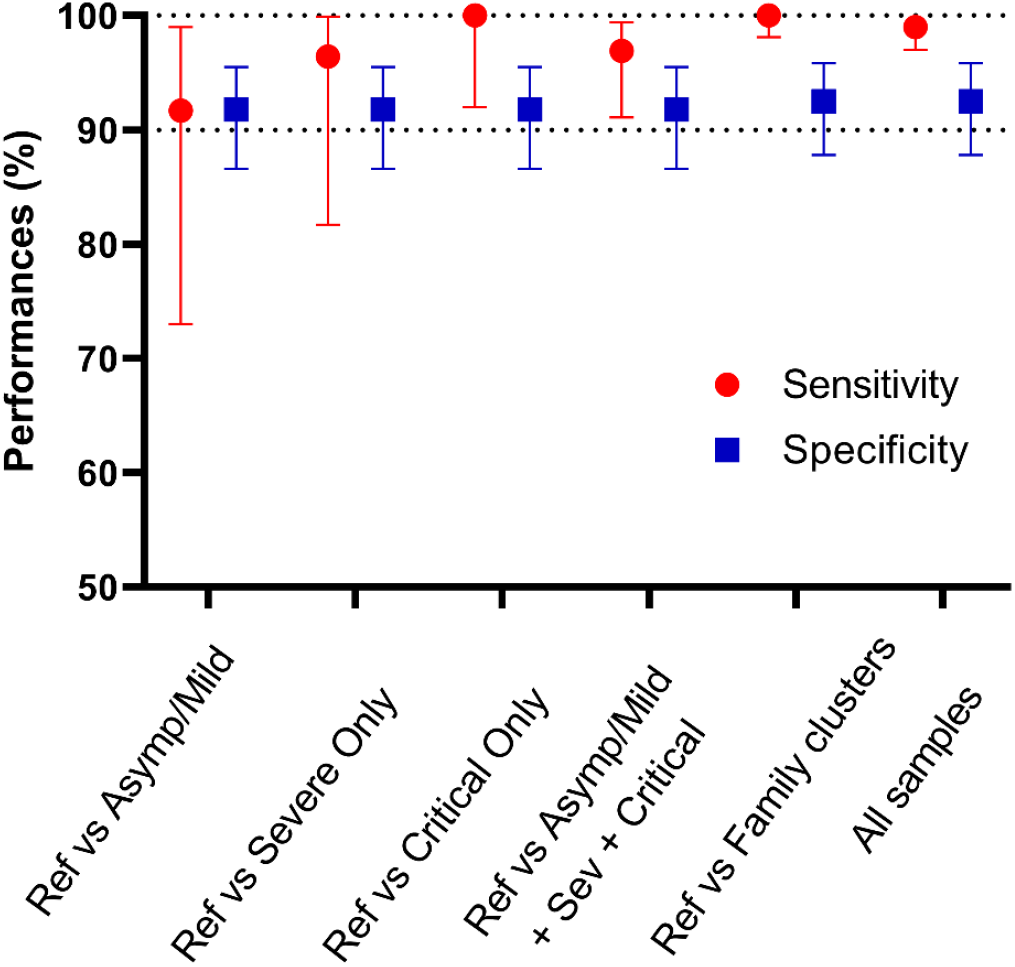
Sensitivities and specificities of anti-SARS-CoV-2 S-RBD IgG, calculated considering only samples collected after 14 days from symptom onset. Different conditions were inspected and compared. Ref group includes SARS-CoV-2 negative samples from pre-pandemic specimens, healthcare workers and patients with rheumatic diseases and HIV.

Although sensitivity and specificity are helpful for clinical purposes, positive and negative predictive values (PPV and NPV) are more relevant in clinical decision making. Using two different scenarios of disease prevalence settings, (a) 4%, as found in a Veneto Region (Italy) survey (13) and (b) 10%, as described in a survey conducted in Geneva (14), PPV and NPV were then estimated. Considering performances derived from sera collected 14 days after the onset of symptoms on Asympt/Mild symptomatic subjects, mimicking a survey conducted in a population not reporting symptoms attributable to COVID-19, the PPV (95%CI) and NPV (95%CI) were 31.8% (21.8%-43.9%) and 99.9% (98.6-100%) with a prevalence of 4%, and 55.4% (42.6%-67.6%) and 99.0% (96.3-99.7%) with a prevalence of 10%.

### 3.5 CLIA results correlation with PRNT _50_ results

Considering all individuals included in the study, a total of 281 leftover samples results of the PRNT_50_ test were available (Table 2 and Supplementary Figure 1). The relationship among the anti-SARS-CoV-2 S-RBD IgG and the corresponding PRNT_50_ titers is shown in Figure 4, panels A and B. Overall, positive associations were found between log_10_ PRNT_50_ titer and log_10_ Ab results. An elevated correlation was obtained (rho = 0.689, p < 0.001) at univariate analyses. At multivariate analyses, performed including Age, Gender and the time from symptom onset and serological determination in the linear regression model, a similar correlation coefficient was found (R^2^ adj = 0.508, rho = 0.712), being only Age (p = 0.013) and time post symptom onset (p = 0.041) statistically significant. In a further sub-analysis, including also disease severity, this latter variable results not significantly associated with log_10_ PRNT_50_ titer.

**Figure 4:**
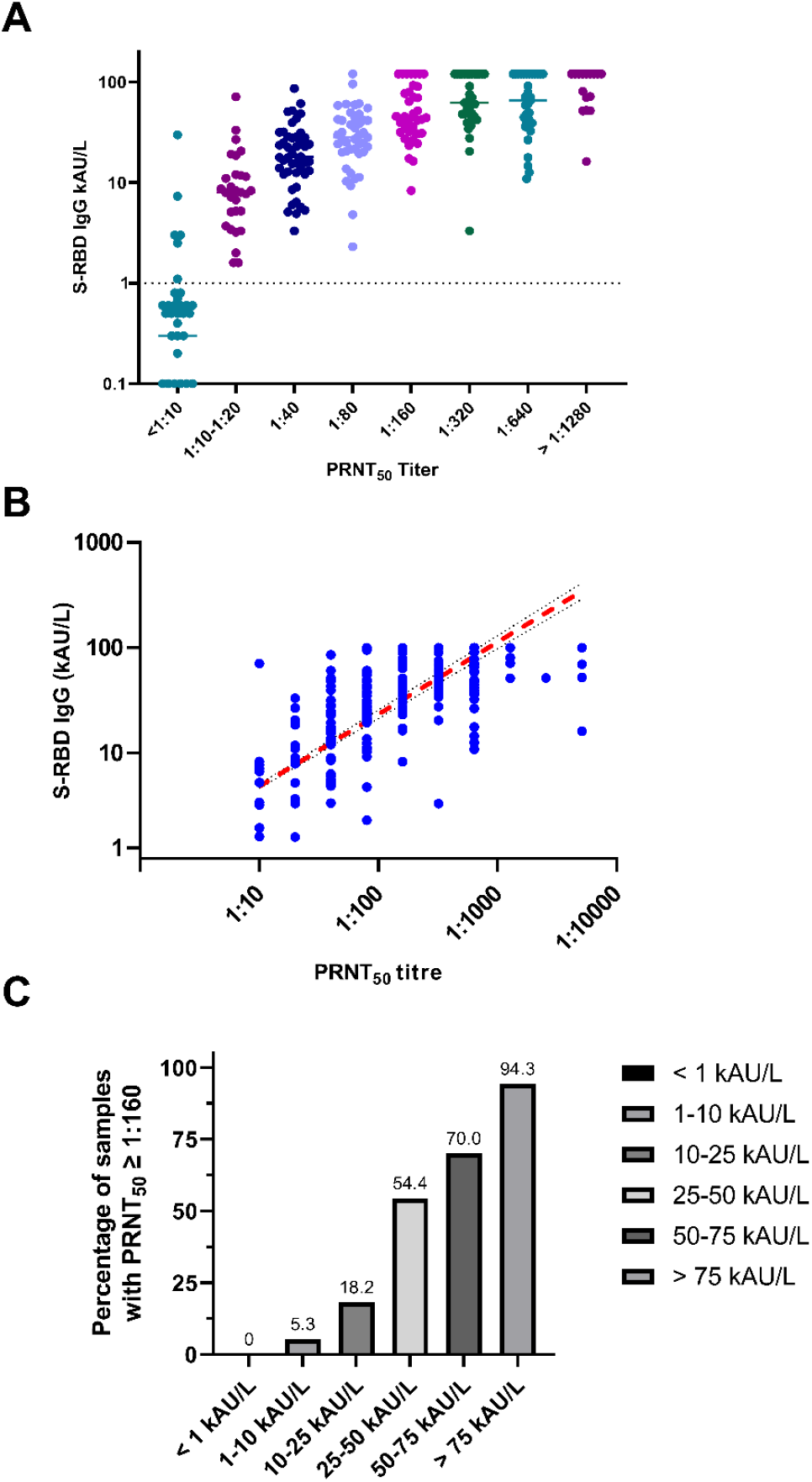
Correlation between the anti-SARS-CoV-2 S-RBD IgG CLIA results and PRNT_50_ titers: A) dot plots presenting the CLIA results with respect to the different PRNT_50_ titers; B) linear correlation of positive PRNT_50_ titers with respect to CLIA results (both in log_10_ scale); C) percentage of samples with PRNT_50_ titers ≥ 1:160 and different ranges of CLIA results.

Since for COVID-19 convalescent plasma treatments a high neutralization titer is advisable, a further analysis was performed (15). Figure 4 (panel C) shows the percentage of samples with a PRNT_50_ titer ≥ 1:160 with respect to the ranges of S-RBD IgG results. For CLIA result above 75 kAU/L, a neutralizing titer ≥ 1:160 was detected in the 94.3% of cases (5% of cases below 1:160 were three samples with PRTN50 equal to 1:80, 1:80 and 1:40).

## 4. Discussion

This paper reports a head-to-head evaluation of the Snibe anti-SARS-CoV-2 S-RBD IgG CLIA analytical performances, since this assay is claimed to be quantitative and the evaluation of these characteristics is especially important for monitoring seroconversion and antibody persistence. Results showed that this assay presents excellent analytical performances, both for precision and linearity. The repeatability was less than 6% for all the studied levels, while intermediate precision was more elevated at the lowest level (1 kAU/L), which is close to the cut-off proposed by the manufacturer (Table 3). Precision performances statistically deviated from the manufacturer’s claims only at the highest level (6.14 kAU/L), as the precision value reported inside the inserts at 5.11 kAU/L were 2.25% and 2.40%.

The adoption of serological testing for monitoring of Ab titers requires, in addition to assay robustness, a good method linearity, to effectively quantify differences between measured values. Our data demonstrate that anti-SARS-CoV-2 S-RBD IgG presents excellent linearity not only within the range of values including the cut-off (0.2-4 kA/L) but also for the highest values (from 5 to 70 kAU/L) (Figure 1); notably, these findings are relevant when considering that, in vaccinated subjects, Ab values above the limit of the method are often detected, requiring a further dilution step for delivering results (data not shown).

On a large panel of blood samples, including pre-pandemic, negative HCW, and negative AI/HIV specimens and SARS-CoV-2 patients with different severity of disease (Asymp/Mild, Severe and Critical), using the pre-defined assay thresholds for calling test results positive or negative, overall sensitivity and specificity were around 97% and 92%, respectively (Figure 3 and Supplementary table 1). The suboptimal specificity is related to the presence of some false-positive results obtained for 14 samples (including 4 pre-pandemic, 2 AI, and 8 HCW specimens). In agreement with the time-dependent nature of antibody response, different results are obtained assessing samples collected at least 14 days post symptoms onset (16). Accordingly, two separate analyses were conducted. In the time frame from 14 days post symptom onset, using all negative subjects as references (Ref), better sensitivity results were achieved for critical rather than severe disease patients, despite the anti-SARS-CoV-2 S-RBD IgG did not differ between the groups of severe and critical patients. Comparing Ref and family clusters, performances of anti-SARS-CoV-2 S-RBD IgG were excellent, being sensitivity 100% and specificity above 92%; remarkably, all samples of this group were collected after 14 days post symptom onset. Considering samples from family clusters, a slight statistically significant time-dependent decrease of anti-SARS-CoV-2 S-RBD IgG was observed, and linear regression allowed to estimate a change in Ab levels, with a confidence of 95%, from −0.17 to −0.04 kAU/L per day (Supplementary Figure 2) and in a further analysis, performed excluding individuals aged < 30 years, findings confirmed the magnitude of the linear slope. These results are fully in accordance with our previously reported data (16), suggesting that, with the exception of some individuals, immunological memory remain persistently elevated for months (17).

The relationship between SARS-CoV-2 antibodies and neutralizing activity remains an essential and open issue. In fact, SARS-CoV-2 neutralizing antibodies (NAb) titer is currently gaining importance for supporting vaccine development, and to aid convalescent plasma therapy. Therefore, due to the high demand for the neutralization test, a surrogate method to evaluate their levels in patients with varying severity of illness at a various time points is strongly advisable, also for circumventing the need to handle live virus in BSL-3 laboratories. For this reason, we assessed the correlation between the plaque reduction neutralization, the gold standard methods for determining the titer of NAb, with anti-SARS-CoV-2 S-RBD IgG levels. Overall, the anti-SARS-CoV-2 S-RBD IgG levels showed a good dynamic range and the response of the method was highly correlated with PRNT_50_ titers (Pearson rho = 0.712 at multivariate analysis) (Figure 4). In addition, when the percentage of samples with a PRNT_50_ titers ≥ 1:160 was calculated with respect to the ranges of anti-SARS-CoV-2 S-RBD IgG, results above 75 kAU/L presented a neutralizing titer ≥ 1:160 in the 94.3% of samples. These results are in accordance with our previously reported findings, performed in different assays, which gave similar results of this anti-SARS-CoV-2 S-RBD IgG. Currently, a small number of studies have validated a range of commercially available SARS-CoV-2 serological assays against a live-virus neutralization test (17–23), and in our knowledge this is the first study comparing Snibe anti-SARS-CoV-2 S-RBD IgG levels and PRTN_50_ titers. Walker et al. evaluated different commercially available assays for their correlation with the microneutralisation assay and reported values ranging from 69% to 100%, with assays measuring total antibodies being the most sensitive (23). Differently, Legros et al. found that Diasorin SARS-CoV-2 S1/S2 kit anti-S IgG titers correlated highly with microneutralization nAb titers (Spearman’s ρ = 0.7075) (22). Patel et al., who evaluated five immunoassays with respect to NAb results, observed that the strongest correlation was rho = 0.81 (with the ELISA from Euroimmun) and the weakest correlation was rho = 0.40 (with Roche CLIA assay) (20).

This study presented several limitations. First, neutralizing antibodies were mainly tested in a well-defined cohort of family cluster, with sera collected at various time points and, therefore, should be confirmed in further studies; second, COVID-19 positive patients were selected retrospectively on the basis of available leftover samples, and third cross-reactivity with seasonal human coronaviruses was not assessed; therefore NPV and PPV could be overestimated. Another limitation of this study is that no longitudinal sera were analyzed and, therefore, we cannot exclude that some patients might have seroconverted at later time points.

In conclusion, the data reported in this study showed that anti-SARS-CoV-2 S-RBD IgG assay achieves excellent analytical and clinical performances. Since specificity results were not 100%, the assay might present a limited number of false-positive results and this characteristic could be further confirmed in a more representative number of samples. However, the correlation with sera neutralization activity was very elevated, demonstrating that the dynamic range of the assay is expanded enough to capture all clinically significant NAb results. Finally, an appropriate threshold could be derived for selecting samples for COVID-19 convalescent plasma therapy.

## Data Availability

Data will be made available upon request

## Acknowledgments

We thank Giulia Vanuzzo for their valuable technical support. We acknowledge Snibe diagnostics for kindly supplying reagents without any influence in study design and data analysis.

## Supplementary Figures

**Supplementary Figure 1:**
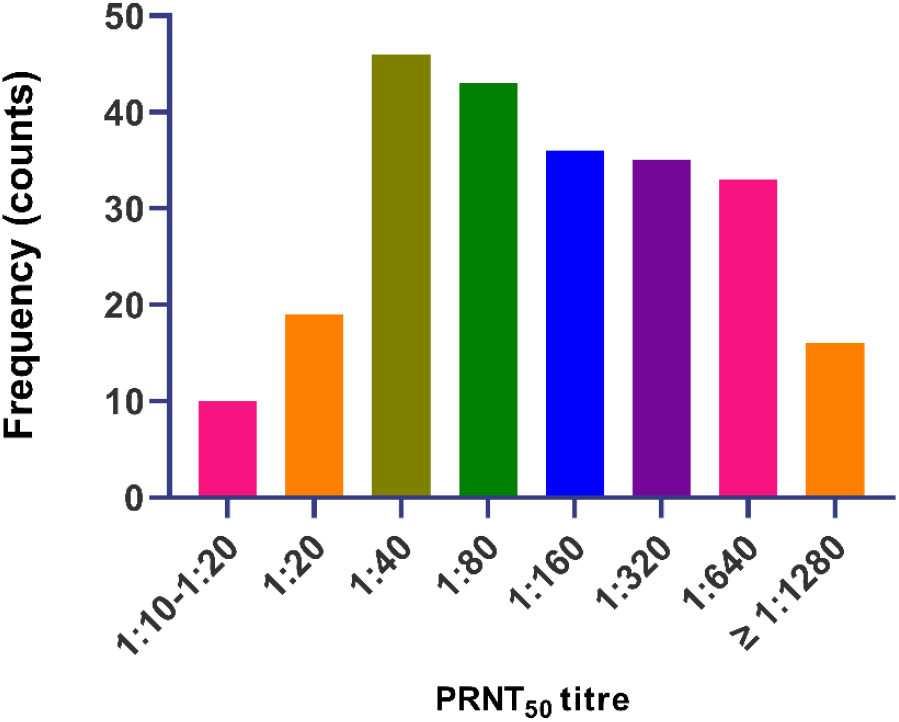
Correlation between the anti-SARS-CoV-2 S-RBD IgG CLIA results and the time from symptom onset on samples from family clusters. This group of individuals was used for evaluating the correlation because: a) samples were collected in a wide interval days; b) it is the most numerous group.

**Supplementary Figure 2:**
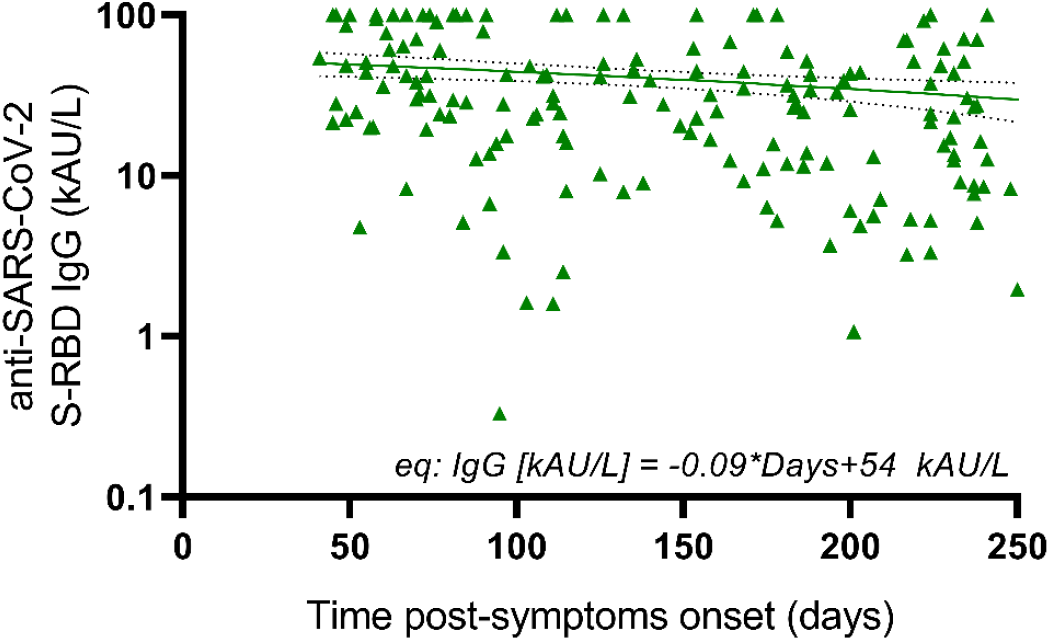
Distribution of the PRNT_50_ titers in PRNT_50_ positive subjects (titer ≥ 1:10).

**Supplementary Table S1:** Clinical performances of anti-SARS-CoV-2 S-RBD IgG, overall and considering only the period ≥ 14 days post symptoms onset. Analyses were repeated by the different groups of individuals.

|  | **Sensitivity**<br>**(95% CI)** |  | **Specificity**<br>**(95%CI)** | **Positive Likelihood ratio**<br>**(95%CI)** |  | **Negative Likelihood ratio**<br>**(95%CI)** |  | **ROC analyses**<br>**(95%CI)** |
| --- | --- | --- | --- | --- | --- | --- | --- | --- |
| | Overall | $\geq 14$ days post<br>symptom onset | Overall | Overall | $\geq 14$ days post<br>symptom onset | Overall | $\geq 14$ days post<br>symptom onset | |
| Ref vs<br>Asymp/Mild<br>Symptomatic | 91.7<br>(73.0-99.0) | 91.7<br>(73.0-99.0) | 91.8<br>(86.6-95.5) | 9.26<br>(5.38-15.93) | 11.20<br>(6.68-18.76) | 0.15<br>(0.10-0.22) | 0.09<br>(0.02-0.34) | 0.917<br>(0.857-0.977) |
| Ref vs Severe | 78.6<br>(67.1-87.5) | 96.4<br>(81.7-99.9) | 91.8<br>(86.6-95.5) | 9.60<br>(5.72-16.09) | 11.78<br>(7.09-19.55) | 0.23<br>(0.15-0.37) | 0.04<br>(0.01-0.27) | 0.852<br>(0.799-0.904) |
| Ref vs Critical | 94.6<br>(86.7-98.5) | 100<br>(92.0-100) | 91.8<br>(86.6-95.5) | 11.55<br>(6.97-19.14) | 11.73<br>(7.16-19.21) | 0.06<br>(0.02-0.15) | 0.01<br>(0.01-0.19) | 0.932<br>(0.898-0.956) |
| Ref vs Severe and<br>Critical patients | 86.8<br>(80.2-91.9) | 98.6<br>(92.5-100) | 91.8<br>(86.6-92.5) | 10.60<br>(6.39-17.59) | 12.04<br>(7.29-19.91) | 0.14<br>(0.09-0.22) | 0.02<br>(0.01-0.11) | 0.893<br>(0.858-0.927) |
| Ref vs<br>Asympt/Mild,<br>Severe and Critical<br>patients | 97.5<br>(81.5-92.1) | 96.9<br>(91.1-99.4) | 91.8<br>(86.6-92.5) | 10.69<br>(6.45-17.71) | 11.83<br>(7.15-19.57) | 0.14<br>(0.09-0.20) | 0.03<br>(0.01-0.10) | 0.896<br>(0.864-0.929)\ |
| Ref vs Family<br>Clusters* | - | 100 (98.1-100) | 92.5<br>(87.8-95.8) | - | 12.92 (7.89-21.2) | - | 0<br>(0 - 0.04) | 0.962<br>(0.943-0.981) |
| All samples | 94.2<br>(91.2-96.3) | 99.0<br>(97.0-99.89) | 92.5<br>(87.8-95.8) | 12.58<br>(7.59-20.83) | 13.22<br>(97.0-99.8) | 0.06<br>(0.04-0.10) | 0.01<br>(0.01-0.03) | 0.933<br>(0.910-0.956) |

\*All samples from Family clusters were collected after 14 days post symptoms onset.

